# KNOWLEDGE AND TREATMENT PRACTICES OF HEPATITIS B INFECTION IN CHILDREN AMONG HEALTH PRACTITIONERS IN KRACHI DISTRICTS IN GHANA-A CROSS-SECTIONAL STUDY

**DOI:** 10.1101/2022.10.28.22281656

**Authors:** Rebecca A. Mpangah, Ernest Akyereko, Gideon K. Acheampong, Patrick K. Nyambah, Michael Ansah-Nyarko, Isaac Owusu, Bismark Sarfo

## Abstract

**Introduction:** Hepatitis B virus (HBV) infection remains one of the neglected infectious diseases. Children infected with HBV are at higher risk of becoming chronic carriers. Barriers to measures against HBV in children is attributed to inadequate knowledge by some health professionals. This study assessed knowledge and treatment practices of HBV in children among health professionals.

**Methods:** A cross sectional survey was conducted among health practitioners (185) in three districts in Krachi using structured questionnaire. Stata version 15 was used to analyze participants’ responses on awareness, knowledge and treatment practices. Pearson’s Product Moment correlation was used to determine the relationship between knowledge, treatment and preventive practices. Multivariate regression analysis assessed the relationships between variables at p<0.05 and 95% confidence interval.

**Results:** 64% of the participants were nurses. 80% were aware of HBV in children and 85% had only fair knowledge about HBV in children. Only 29% indicated good knowledge and management practices of HBV in children. There was a positive relationship between knowledge, and treatment (*r (183) = .67, p < .001)* and preventive (*r (183) = .54, p < .001*) practices. A unit increase in awareness of HBV in children leads to 1.42 units increase in knowledge *(p < .01, 95% CI; .543, 2*.*296)*, while a unit increase in knowledge result in 1.3 units increase in treatment (*(p < .01, 95% CI;.912, 1.680*) practice of HBV in children.

**Conclusion:** Participants demonstrated only fair knowledge about HBV in children. Seminars and workshops on HBV in children for health professionals must intensify.

## Introduction

The estimated prevalence of hepatitis B virus (HBV) in Ghana among adolescent population is 14.30%, and 8.36% among adult with 0.55% in children under five years [1]. The overall prevalence of HBV in Ghana is estimated at 12.3% [2]. This makes the country highly endemic to HBV [3].The prevalence in rural areas is reported to be 13.3% whiles that of urban areas was reported to be 12.2% [2]. Approximately 15% of the Ghanaian adult population are living with HBV of which 90% was acquired through mother-to-child transmission (MTCT) [2]. Though there have been vaccines and treatment therapies such as use of antiviral drugs endorsed by the World Health Organization including nucleotides analogues, entecavir, tenofovir and lamivudine for the management and control of HBV, there still remain a larger number of persons living with HBV untreated which exposes them to developing into chronic carriers and increases the burden on the nation [2]. Whereas about 70% of HBV infected persons within the Americas and Western pacific regions have been treated successfully, only 10% got access and received treatment for HBV from the WHO African region [4].

The barriers to the effective administration of vaccines and other preventive measures is attributed to inadequate knowledge about HBV by persons responsible for the active implementation of the elimination policy set by WHO as well as lack of resources, vaccines, protocols and logistics and cost involved for the effective implementation of good preventive practices [5]. Despite a decrease in the incidence of HBV, prevalence remains higher due to insufficient coverage rates of vaccination and other limitations in other preventive measure [6].

Healthcare providers who are to see to the implementation policies towards the eradication of HBV appear to be uncertain about the measures to be implemented and how to go about it as it is found that some facilities in Ghana do not administer the “at birth” dose of the vaccine as required [7]. Reports indicate that, most medical practitioners as well as students have been found to have received no formal training on the disease and how it should be managed [8,9]. The birth dose of the HBV vaccine is supposed to be administered within 24 hours after birth to neonates born to HBV positive mothers. However, studies found that, they are delayed until the routine vaccination periods for children in Ghana which is 6, 10 and 14 weeks after birth, before the vaccine is administered when the child might have already acquired the virus [7].

Due to the asymptomatic nature of the virus, children are at higher risk of developing into chronic carriers before it is diagnosed and they are also prone to getting the virus horizontally at school through open cuts and scratches since the virus can survive for at least seven days on environmental surfaces [10]. This as a result makes children more vulnerable to HBV disease.

There is very limited studies conducted in the area of knowledge of health practitioners in Ghana to assess why some medical practitioners do not administer the required dose of vaccination in children born to HBV positive mothers especially in rural areas for that matter. This study assessed health practitioners’ knowledge and treatment of HBV in children in the Krachi districts which falls within the rural and peri-urban communities in Ghana where prevalence is about 13.3% as against 12.2% in the urban areas [2].

## Methodology

### Study Area and Population

The setting for this study included health facilities within the Krachi districts namely Krachi-Nchumuru, Krachi East and Krachi West. According to the 2021 Population and Housing census, the respective populations of these districts are 79,934, 116,804 and 61,128 [11].

The Krachi-Nchumuru district currently does not have a Government Hospital. However, there are both private and public health service providers. There are 5 Health Centers, 2 Mission Clinics, 9 Maternal and Child Health/ Family Planning (MCH/FP) clinics. For this study, participants were enrolled from the St. Luke Health Center at Chinderi (Private), Borae Health Center (Public) and Banda Health Center (Public).

The Krachi East District has 8 private and public health centers. Participants were enrolled from the Dambai Health Center (Public) which is in the district capital.

Krachi West district has 9 private clinics and 8 public health centers and 1 district hospital.

The district hospital serves as the main referral center within the district where the other two neighboring districts refer their patients for better treatment in case they need extra healthcare services that are not in their facilities. The participants for this study were enrolled from the district hospital.

The selected districts for this study are combination of rural and peri-urban areas, hence the main occupations of the people are farming, fishing and trading. The districts were chosen for the study because they give a reflection of both the rural and peri-urban areas which are mostly neglected in terms of health facilities and research, although they equally carry a lot of disease burden in the country.

### Study design

The study adopted the quantitative approach using the cross-sectional survey design which was conducted between June and July in 2019.

The participants for the study are health practitioners who are practicing in the selected health facilities in the three Districts including Doctors, Nurses, Midwives, Physician Assistants, Community Health workers and all other Public Health practitioners within the health units who see to the well-being of patients.

### Sample Size

The single population proportion formula below was used to estimate the sample size:

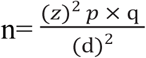

Where;

**n=** the desired sample size

**z**= confidence interval at 95% set at 1.96

**p** =prevalence (12.7%) of knowledge among health workers that HBV vaccine can prevent the transmission of the virus to newborns of HBV infected mothers in the Eastern region of Ghana (Adjei et al. (2016)

**q**= the difference between 1 and p

**d**= maximum error allowed (5%, if the confidence level is 95%); 0.05. Substituting into the formula, the sample size was estimated as follows;

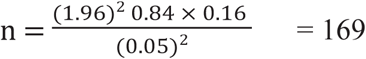

#### Sampling technique

Health workers available within each of the selected facility and who were willing to participate in the study were enrolled. Participants were proportionally sampled from the three districts for the estimated sample size.

### Inclusion criteria

Health practitioners including Physicians, Nurses, Midwives and other Public Health professionals within the health centers who have had at least six months working experience after training and have consented were enrolled into the study.

### Exclusion criteria

Participants who did not consent and also those who were still undergoing training or internships or had less than six months working experience in the health profession were excluded from the study.

### Study variables

*The variables for this study were categorized into three: i) demographic characteristics which included sex, age, work experience, healthcare group and educational level, ii) knowledge and awareness of HBV infection in children iii) treatment/management and prevention of HBV infection in children*.

#### Study questionnaire

The questionnaire for this study was adapted from previous studies on awareness, knowledge, treatment/management and prevention practices of HBV among health care workers [2,12-14]. The questionnaire has 38 items aside the demographic information. The questionnaire comes with five main sections (section A to E). Section A consists of introduction and ethical issues of the study; section B is about the demographic characteristics of participants; Section C measures awareness level; section D measures knowledge about hepatitis B in children and contains 22 items; section E measures treatment/management and prevention practices with a sub section that measured sources of knowledge and availability of management resources which also contain 16 items in all.

### Scoring of participants responses on awareness, knowledge and treatment and prevention practices of HBV

Each correct response was scored one (1) and incorrect responses zero (0). Awareness was scored in two different categories of general awareness scored as correct and incorrect and awareness about hepatitis B in children which were also scored accordingly. On knowledge score, less than 7 correct responses represented poor knowledge, 7-16 correct responses represented fair knowledge and 17 -22 correct responses represented good knowledge. This was based on the normal distribution curve. Concerning the treatment, 3 or less correct responses represented low treatment practices while 4 or more correct responses represented good treatment practices. Prevention was also scored based on individual items or practices as correct and incorrect.

### Data analysis

Data management and analysis was performed using Stata software version 15.0. Data were assessed for normality for some of the selected variables to ensure that it was not skewed before subsequent analysis was performed. Frequency distribution was used to analyze demographic characteristics as well as responses by participants on awareness, knowledge and treatment and preventive practices of HBV in children. After scoring participants knowledge on HBV in children on a 22 question items, the scores were further categorized into three knowledge levels (poor, fair and good) through the use of median split format of categorization. If participants correctly responded to only 7 items and below, it was categorized as poor knowledge; correct responses for items 7 to 16 was categorized as fair knowledge and those who correctly answered 17 or more knowledge items were classified as having good knowledge of HBV transmission in children.

Similarly, a composite score on management and prevention practices was generated and categorized into poor or inadequate practices (3 or less correct responses) and good treatment practices (4 and greater correct scores).

The Pearson’s Product Moment Correlation which estimates whether there are relationships between variables, and the strength and direction of this relationship, was used to determine the relationship between knowledge, treatment and prevention and other variables like source of knowledge and awareness. Finally, the multivariate regression analysis was performed to assess the relationships between knowledge about hepatitis B in children among health practitioners and other variables at 0.05 significance level with 95% confidence interval.

### Ethical approval

This study received ethical clearance from the Ghana Health Service Ethics Review Committee (GHS-ERC 022/01/19) and permission was granted by the participating health facilities before the commencement of the study.

## Results

The detailed socio-demographic characteristics and source of knowledge of study participants is presented in Table 1. A total of 185 health workers participated in the study out of which 90(48.65%) were males and 95(51.35%) were females. With participants’ age, majority (66%) of them were within 26-35years, and very few (1.6%) were between 46-55years. Most of them (43.78%) had 2 to 5 years of work experience and only 1.08% had over 15years work experience.

Majority of the participants (156/185, 84.32%) had certificate and diploma qualification. Only 3 Doctors (1.62%) were available for participation in this study. The ‘Others’ representing Public Health Officers, Community Health Nurses and Disease Control Officers were only 9.19% of the study participants (Table 1).

**Table 1.** Socio-demographic characteristics and source of knowledge of study participants

General awareness of HBV among the health professionals and awareness of HBV in children specifically were assessed (Table 2).

**Table 2.** General awareness of Hepatitis B virus among the participants

### Awareness about HBV

Although all the 185(100%) participants in the study were aware of HBV infection, 20% (37) were not aware of HBV infection in children (Table 2).

Subsequent to testing the awareness level of HBV among the participants, their knowledge of the virus in children was tested and their responses were scored as correct or incorrect. In all, 22 knowledge items were scored as summarized in Table 3.

**Table 3.** Descriptive statistics for knowledge of hepatitis B in children among the participants

The results from Table 3 show that, with knowledge about hepatitis B virus, 91.9% (170) of the participants correctly identified HBV as a virus as well as its ability to be transmitted through contaminated/infected blood transfusion. Also, 132(71.35%) answered correctly on HBV horizontal transmission from child to child through bodily fluid contact, 130(70.27%) identified the availability of vaccine for prevention of HBV while 129(69.73%) also correctly answered that immune response test is necessary after vaccination. Though majority of them provided correct answers to the questions, there were some responses that raise lots of questions. In all, there were incorrect responses to each of the questions that tested their knowledge on HBV as demonstrated in Table 3. For instance, 114 of the participants failed to identify correctly that, immunoglobulin helps in HB prevention after few hours of exposure. Though 66.7% (123) knew that children are at higher risk of becoming chronic carriers than adults, 61.62% (114) were unable to identify that adults are more likely to recover than children. Mother-to-child transmission at birth was correctly answered by 144 (77.8%) but 129 (69.7%) were not able to identify that, HBV cannot be transferred to a fetus. Liver cancer and cirrhosis have been correctly identified to be linked with HBV. Furthermore, 118(63.8%) of the participants failed to correctly identify that, the HB virus can survive on surfaces for over 24 hours and 131(70.8%) wrongly identified HBV as symptomatic within the first 6 months.

After the individual item analysis, the scores of each knowledge item were summed up for a composite knowledge score. These scores were further categorized into three knowledge levels-*Poor, Fair* and *Good*, through the use of median split format of categorization. Summary of this analysis of knowledge levels of HBV is shown in Table 4.

**Table 4.** Summary statistics for knowledge level of HBV in children

Majority (85.4%) of the participants had fair knowledge on HBV in children, with only 5.4% having good knowledge of HBV. About 9% (17/185) had poor knowledge of HBV in children.

Further to assessing the knowledge levels of HBV, the participants were assessed on the treatment/management and prevention aspects of HBV in children on 7 items which were scored as correct or incorrect as shown in Table 5.

**Table 5.** Descriptive analysis for scores on management and prevention items of hepatitis B in children

The management and prevention practices of HBV in children was assessed among participants and 157(84.86%) correctly identify the need for screening pregnant women for HBV before they give birth. However, 114(61.08%) participants incorrectly responded that they would not vaccinate children against HBV irrespective of their status or that of the mother. One hundred and thirteen (113) health workers also incorrectly answered that, they have not adequately treated HBV in a child before while 118 (63.78%) rightly said they will not wait for treatment of HBV to be done later while attending to other conditions.

A composite score on management and prevention practices was generated and categorized into poor or inadequate practices (3 or less correct responses) and good treatment practices (4 and greater correct scores) as shown in Table 6.

**Table 6.** Descriptive statistics for treatment practices of HBV in children

Table 6 demonstrates that only 29.2% (54/185) indicated good knowledge in the management and prevention practices of HBV in children. Majority of the participants (70.8%) had poor or inadequate knowledge of HBV treatment practices in children.

Participants were further asked about the availability of resources for management and prevention of HBV in their facilities and their responses are summarized in Table 7.

**Table 7.** Descriptive statistics for resources availability for management and prevention of *HBV*

Table 7 shows that, 53.5 % (99/185) of the participants reported that there are no treatment protocols at their facilities to manage HBV while 129 (69.7%) also agreed that no drugs for the management of HBV are available. Though 60.5% (112) agreed that there are vaccines available at their facilities, 63.8% (118) reported that no vaccination programs have been organized by the Ministry of Health in their area.

The correlation between knowledge of HBV and management and prevention practices adopted by health practitioners in the district was further estimated (Table 8) using the coefficients (r) of the Pearson’s Product Moment Correlation.

**Table 8.** Summary statistics for the relationships between knowledge about hepatitis B in children among health practitioners and management and prevention and other variables

Table 8 shows that there was a strong positive relationship (*r (183) = .67, p < .001)* between knowledge and management practice of HBV in children by the study participants. Also knowledge has a strong positive relationship (*r (183) = .54, p < .001*) with prevention practices among the study participants.

Meanwhile there was a moderate positive relationship between knowledge and sources of knowledge (*r(183) = .44, p < .001*) and also between knowledge and resource availability (*r(183) = .32, p < .001*) among the study participants as far as hepatitis B in children is concern.

The relationship between knowledge and awareness of hepatitis B among the health practitioners is weak although it is positive (*r(183) = .25, p < .001*).

Further analysis was conducted to determine the association between variables while adjusting for possible confounders using the multiple regression analysis.

After adjusting for possible confounders such as age, sex, educational level, and work experience, the following variables had significant positive association with knowledge; awareness, treatment, preventive practices and source of knowledge (Table 9). From Table 9, it can be inferred that a unit increase in awareness will lead to 1.42 units increase in knowledge *(p < .01, 95% CI; .543, 2*.*296)*. While, a unit increase in knowledge will result in 1.3 units increase in treatment (*(p < .01, 95% CI;*.*912, 1*.*680*), 0.78 unit increase in prevention (*p < .01, 95% CI382, 1*.*181)* and 0.62 unit increase in source of knowledge (*(p < .05, 95% CI:074, 1*.*69)* (Table 9).

**Table 9.** Summary statistics of multiple regression analysis for relationships between knowledge about hepatitis B in children among health practitioners and other variables

## Discussion

This study has demonstrated that although all the participants (Doctors, Nurses, Midwives, Physician Assistants, and other Public Health professionals) were aware of HBV infection in the general population, some of them were not aware that HBV can infect children. Majority of the participants had demonstrated only a fair knowledge of HBV in children, with only few having good knowledge of HBV in children. Some of them had poor knowledge of HBV in children.

Furthermore, only few of the participants showed good knowledge in the management and prevention practices of HBV in children. Majority of them had inadequate knowledge in the management and preventive practices of HBV in children.

In terms of availability of resources for management and prevention of HBV, about half of the participants indicated that there were no treatment protocols at their facilities to manage HBV, while majority of them reported that there were no available drugs for the management of HBV.

Meanwhile some agreed that there are HBV vaccines available at their facilities, while others reported that no HBV vaccination programs have been organized by the Ministry of Health in their area.

The awareness level of HBV in the general population by the participants is higher than the moderate awareness level reported in Cameroon, Southern India, and Saudi Arabia [12,15-16]. This general high level awareness of HBV among the participants is a good indication of first level achievement when it comes to infectious diseases transmission and public health issues among these health practitioners. This awareness level could be linked with training that they obtained from school before practice, since majority of them indicated that they have had training on HBV in school. This finding is consistent with the similar awareness level reported by Adjei et al. (2016) [2] in a study conducted in the Eastern region of Ghana where awareness campaigns that are organized in the country through various media platforms and screening programs contributed to this high level of HBV awareness among the health professionals.

Against the background that some of the participants are not aware of HBV in children, it raises issues of public health concern. This is because HBV infection and infectivity rate among children is high since they are vulnerable in relation to the exposure and progression of disease in adult. Mitchell and colleagues (2010) [17] reported that most of the factors that hinder the prevention and control of HBV is the lack of inadequate awareness and knowledge of healthcare providers. Therefore awareness of HBV in children should be intensified as they have 85 to 90 per cent chance of developing into chronic carriers [18, 4] compared with adults. Awareness campaigns can be embedded in vaccination programs, HIV educational campaigns among others.

Knowledge about a disease or condition serves as a gate way or an opening step towards modification of desirable behavior [19]. In general, this study showed fair knowledge of health practitioners on HBV in children, which is consistent with moderate knowledge level reported in a study conducted in Jhalawar in India [20]. It is however not consistent with another study conducted in Lagos in Nigeria which reported higher or good knowledge of HBV among health professionals [19]. The report from this study is better than the low levels of knowledge recorded in Sharourah in Saudi Arabia among health workers [21]. The disparity in the knowledge level in this study compared with the others could be associated with the differences in the study areas. Whereas the current study was conducted in rural and peri-urban areas, the other studies were conducted in urban centers.

Although this study was conducted in rural and peri-urban areas in the Krachi Districts, the knowledge level of HBV among the participants is better than what was reported in another study in the Eastern region of Ghana which is an urban area where knowledge level is expected to be high [2]. Knowledge about mother to child transmission at birth was good among the participants and this is consistent with other studies [22] in Guangdong in China among doctors and nurses in rural and urban health facilities. Also, higher knowledge score items were recorded by respondents for infected blood transfusion and horizontal transmission from child to child through bodily fluid contact with infected persons, however there was misunderstanding about transmission from mother to her fetus by most health practitioners in the study. Two-thirds of the health practitioners did not know that HBV cannot be transmitted to a fetus in the womb unless there is bodily fluid contact from infected persons to an infant at birth. Such knowledge gaps need to be addressed. In addition, a greater number of the health workers in the study did not know that, jaundice is a symptom of hepatitis B viral infection in its chronic stage. Other report showed that 25% of all chronic carriers of HBV develop hepatocellular carcinoma or cirrhosis of the liver [23]. In this study, over 50% of the health practitioners know about the association between HBV and liver cancer but a lesser percentage knew HBV as a major cause of liver cancer which is consistent with the findings from another study in Nigeria and India [20, 24]. The lapses in the knowledge of symptoms of HBV in children among the participants could be linked to the low levels of in-service trainings that are organized in the institutions as part of the health system strengthening to upgrade knowledge acquisition of health practitioners after their school training. New things are discovered each day but if this knowledge are not conveyed to the right people who need it for action as in the case of practitioners, there will always be misconceptions and poor practices leading to poor health outcomes. Hence there is the need for improved knowledge in HBV among the health professionals in the study area. This could be achieved through seminars and various awareness campaigns across the districts concerning hepatitis B viral infection. Therefore, the need to intensify programs and health promotion strategies including education for practitioners as well as the general public about HBV in children cannot be overemphasized. When good knowledge or information on the mode of transmission is known it can translate into breaking transmission chain and lead to reducing the burden of HBV especially among children.

Concerning prevention practices of HBV in children, this study has shown that, majority of the health practitioners are more likely to practice preventive care such as screening of pregnant women before delivery to ensure safety and prevention of mother to child transmission. This however, is in contrast to another study by Adekanle and colleagues (2015) [24] in Nigeria which observed that prevention practices were poor among health practitioners though the study was carried out in an urban setting. Other prevention practices such as administration of vaccine with immunoglobulin and at birth dose of the HBV vaccine to babies within 24 hours after birth is reported to be practiced by most of the participants in the current study. Insufficient supply of vaccines or the lack of it in health facilities could hinder the administration of this vaccine by majority of these health professionals to prevent mother to child transmission of HBV.

Surprisingly, majority of the study participants had no knowledge of the asymptomatic nature of HBV in children within the first six months. This implies that, health practitioners may be looking forward to seeing symptoms of the HBV before they try to take control measures which could lead to too many missed opportunities. This gap in knowledge may be as a result of unavailability of periodic training to validate the knowledge of health practitioners on new information about the disease as well as the lack of treatment protocol. Both the availability of the vaccines and the presence of skilled professionals like midwives with adequate knowledge on HBV in children will help in the control and elimination process of the virus [25].

In terms of hepatitis B treatment, about half of the participants indicated that they prioritized treatment for adults presenting with HBV over children with the misconception that there is more risk associated with adults than with children. Also majority of them indicated that they have not adequately treated a child with HBV before. This is a clear demonstration that, more attention is given to adults than children with HBV treatment due to gap in knowledge and lack of experience with HBV in children. Meanwhile some of the health practitioners indicated that they tend to give priority in treating other diseases than HBV. This again shows that some of these practitioners lack adequate knowledge about the burden of HBV especially with the outcome of opportunistic diseases like liver cancer or cirrhosis. Opportunities must be created for these health practitioners to attend seminars and workshops on hepatitis B because in-service training increases ones knowledge as well as good and better practices [26].

This study has demonstrated a significant and positive relationship between knowledge about HBV in children and treatment or management practices. Similar findings were reported by Liu and colleagues (2018) [27] in three larger cities in China among health workers. The correlation between knowledge and HBV management indicates that, as knowledge levels increase, treatment and management practices improve significantly and vice versa.

There was also, a positive relationship between knowledge and prevention practices in this study. Indeed, prevention practices increase with increasing knowledge levels. This implies that, once knowledge of health practitioners concerning hepatitis B virus infection is improved, it will lead to a proportional increase in preventive practices to protect children against HBV and decrease morbidity and mortality. The findings is consistent with a study conducted in Guangdong in China where knowledge was found to be related to prevention of mother to child transmission among rural and urban doctors and nurses [22].

In light of the observations from this study, the World Health Organization’s target of achieving 90% vaccination coverage against HBV by 2030, must first focus on achieving higher knowledge, preventive and management practices among health practitioners especially those in the rural areas to help improve on their services.

Non-governmental Organizations (NGOs) and other governmental agencies should provide educational and training support in addition to organizing seminars and workshops on HBV transmission in children to health professionals.

## Limitations of the study

Being a cross-sectional study, the findings cannot be generalized to health professionals in other districts in Ghana. The study also could not delineate awareness, knowledge and management practices in one category of health professionals, example Doctors from other professionals for targeted future education and training.

## Conclusion

This study concludes that some health professionals in the study area are still not aware of HBV in children and that in all, the participants’ demonstrated only fair knowledge about HBV in children. There are still lapses in certain key knowledge items about HBV in children which might influence treatment and prevention practices. This translated into how some of them prioritized HBV management in adult over children. The study further shows significant and positive relationship between knowledge about HBV in children and treatment and preventive practices by health professionals. Hence the need to continue organizing seminars and workshops and in-service trainings on HBV in children for health professionals in rural and peri-urban areas cannot be overemphasized.

**Figure 1:**
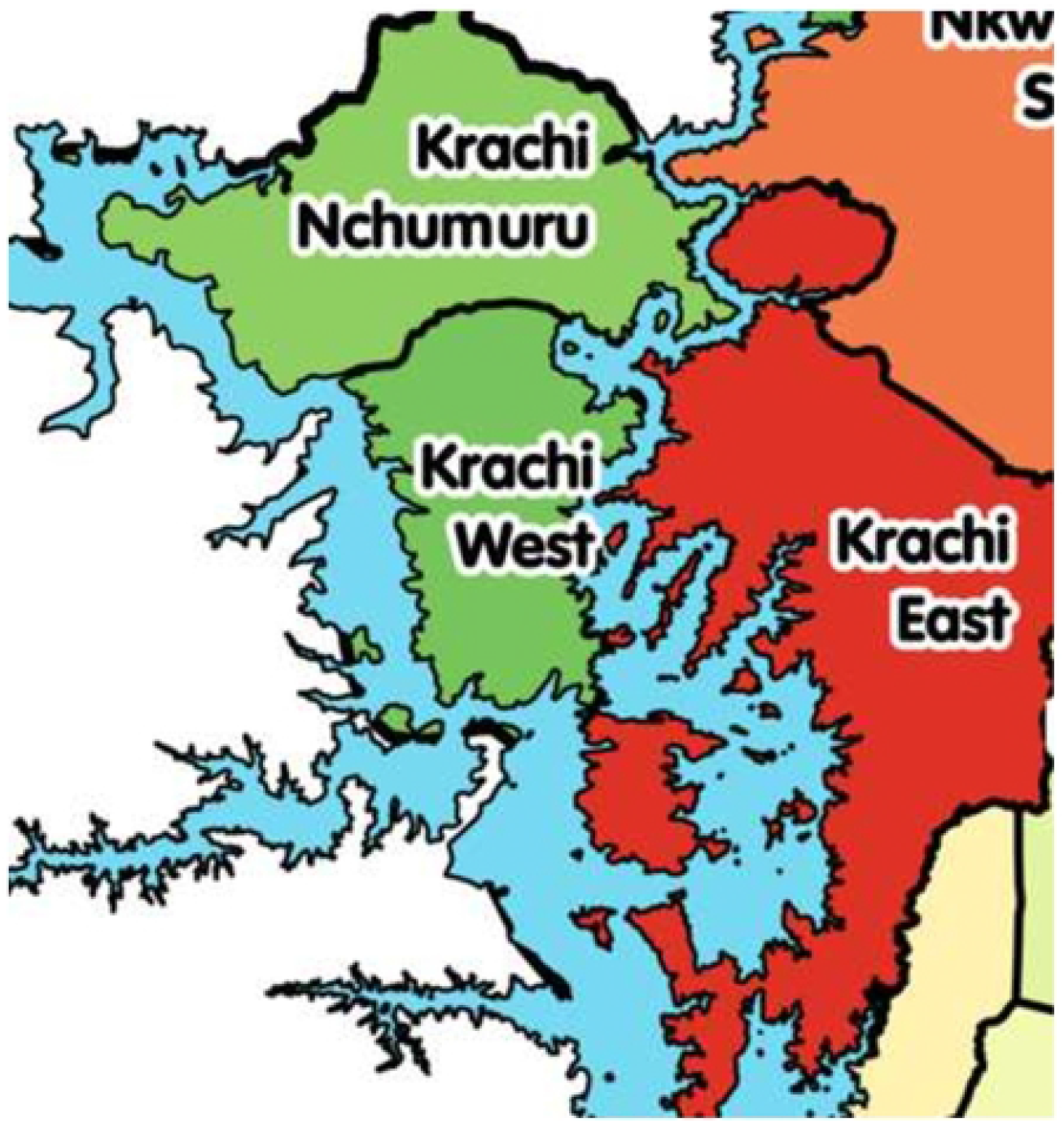
A map showing the three districts namely, Krachi Nchumuru, Krachi West and Krachi East where the health professionals were drawn from

## Data Availability

Data cannot be shared publicly because the data is still being analyzed for additional manuscript

## Authors’ contributions

RAM made contributions to study conception, acquisition of data and analysis and drafting of the manuscript. EA, GKA, PKN, MAN and IO contributed to data analysis and interpretation, and drafting of the manuscript. BS contributed to study conception, design, drafting of the manuscript, interpretation of the data and critically reviewed the manuscript for intellectual content. All authors read and approved the final manuscript.

## Acknowledgements

Many thanks go to the management of the health facilities who participated in the study.

## Abbreviations

HBV: Hepatitis B virus
HI: Human Immunodeficiency Virus
MCH/FP: Maternal and Child Health/ Family Planning
MTCT: Mother-to-Child Transmission
WHO: World Health Organization.

## Consent for publication

Consent and permission were obtained for the publication of these data from the participants and the health facilities.

## Availability of data

The datasets used and/or analysed during the current study are available from the corresponding author on reasonable request.

## Additional files

Cover letter.

